# The neurocircuitry of cue-induced cannabis craving in Cannabis Use Disorder: A functional neuroimaging study

**DOI:** 10.1101/2025.04.03.25323289

**Authors:** Valentina Lorenzetti, Hannah Sehl, Arush Honnedevasthana Arun, Eugene McTavish, Adam Clemente, Hannah Thomson, Marianna Quinones-Valera, Alexandra Gaillard, Emillie Beyer, Diny Thomson, Janna Cousijn, Izelle Labuschagne, Peter Rendell, Gill Terrett, Chao Suo, Lisa-Marie Greenwood, Victoria Manning, Govinda Poudel

**Affiliations:** Neuroscience of Addiction and Mental Health Program, Healthy Brain and Mind Research Centre, School of Behavioural and Health Sciences, Faculty of Health Sciences, Australian Catholic University, Fitzroy, Victoria, Australia; Addiction and Clinical Neurosciences Laboratory, Department of Health Sciences and Biostatistics, Swinburne University of Technology, Hawthorn, Victoria, Australia; Turner Institute for Brain and Mental Health, School of Psychological Sciences, Monash University, Clayton, Victoria, Australia; Neuroscience of Addiction Lab, Center for Substance Use and Addiction Research (CESAR), Department of Psychology, Education & Child Studies, Erasmus University Rotterdam, The Netherlands; School of Behavioural & Health Sciences, Australian Catholic University, Fitzroy, Victoria, Australia; School of Psychology, University of Queensland, Brisbane, Queensland, Australia; School of Medicine and Psychology, Australian National University College of Health and Medicine, The Australian National University, Canberra, Australia; Monash Addiction Research Centre, Eastern Health Clinical School, Monash University, Box Hill, 3128, Victoria, Australia; Turning Point, Eastern Health, Monash University, Melbourne, VIC, Australia; Mary MacKillop Institute for Health Research, Australian Catholic University, Melbourne, Victoria, Australia; Braincast Neurotechnologies, Melbourne, Victoria, Australia

**Keywords:** cannabis, cue-reactivity, craving, fMRI, cannabis use disorder, neuroimaging

## Abstract

**Background:** A common feature of Cannabis use disorder (CUD) is an intense reactivity to cannabis cues, which are becoming increasingly visible due to growth in the decriminalization, accessibility and marketing of cannabis products. The brain’s automatic reactivity to cannabis cues can trigger craving and subsequent use. This study aimed to test neural activity during cannabis cue-induced craving in non-treatment seeking individuals with moderate-to-severe CUD, with past attempts to cut down/quit.

**Methods:** The study examined 65 individuals with moderate-to-severe CUD and 43 controls, with a fMRI cannabis cue-induced craving task and assessment of mental health, substance use, and cognitive testing. Group differences in neural cue-induced craving were examined, adjusting for age and sex; correlations with cannabis use characteristics were assessed, accounting for recent substance use.

**Results:** Individuals with a CUD relative to controls showed greater brain activity during cannabis cue-induced craving in the superior/middle occipital, medial/lateral OFC, anterior/posterior cingulate, cerebellar, hippocampus, middle temporal and lateral parietal cortices (*p* < .05; cluster *k* > 10, FWE-corrected). Greater occipital/cerebellar activity correlated with greater subjective arousal towards cannabis images and cannabis withdrawal scores, while anterior cingulate/inferior parietal activity negatively correlated with urinary level of 11-Nor-9-carboxy-Δ^9^-tetrahydrocannabinol:creatinine (*p’s*<.05).

**Conclusions:** Exposure to cannabis cues can elicit greater activity within salience evaluation/attention, motivation and disinhibition pathways of addiction neurocircuitry in people with moderate-to-severe CUD, consistent with prominent neuroscientific theories of addiction and findings with other substances. Interventions which can suppress brain activity in salience and attention circuits during cannabis-induced craving may help reduce craving and subsequent use.

## 1 Introduction

Around 22 million people globally experience a cannabis use disorder (CUD) during their lifetime, representing around one in four who use cannabis (1). Since 2019, there has been a 32% increase in the incidence of CUD and a 39% increase in CUD-related disability-adjusted life years concomitant with accidental poisoning, schizophrenia, anxiety, depression, and road traffic injuries (2). CUD can be associated with significant adverse psychosocial outcomes, such as withdrawal, and a higher prevalence of anxiety, depression, and psychotic disorders (3). Interestingly however, only a minority (∼13 %) of individuals with a CUD been reported to seek treatment (4). Most people with CUD attempt to reduce or cease use without formal treatment, though research suggests unsuccessful attempts are common, with one study showing 61% make subsequent attempts within 2-3 months (5). These statistics are concerning and highlight the need to uncover the mechanisms underlying CUD in individuals who continue consuming cannabis despite attempts to reduce or cease use.

A core feature contributing to continued cannabis use, relapse or failed attempts to reduce cannabis use, and predicting heavier use in CUD is the experience of strong craving – an intense desire/preoccupation to use cannabis, which may be exacerbated after being exposed to cannabis-related cues (e.g., environmental cues such as images of cannabis, paraphernalia in a shop window, smelling the odor of cannabis at social gatherings, or internal sensations such as stress) (6). As more jurisdictions decriminalize cannabis products globally, cannabis-related cues in the environment will become increasingly ubiquitous, triggering cravings and making it difficult for people wanting to reduce or quit using cannabis. Cannabis cue-induced craving has been ascribed to alteration of brain function within selected brain reward pathways (e.g., striatum, prefrontal regions [anterior cingulate, middle frontal], parietal regions (posterior cingulate/precuneus) and additional brain areas (7).

Functional magnetic resonance imaging (fMRI) studies have examined brain activity in cannabis users during fMRI tasks using cannabis cue-induced craving [(8, 9); for a systematic review, see (10)]. These studies have shown increased brain activity in cannabis users during exposure to cannabis cues in regions involved in cognitive processes typically altered in CUD [(8–10). Such cognitive domains and their underlying brain regions include: reward processing (e.g., nucleus accumbens); habit formation/learning (e.g., striatum, hippocampus); motivation and disinhibition (e.g., prefrontal cortex [PFC], orbitofrontal cortex [OFC], frontal medial cortex); self-monitoring and awareness of environmental stimuli (e.g., precuneus) [(8, 9, 11); and salience pathways (e.g., occipital regions, posterior cingulate cortex [PCC]) [(8, 9, 12).

Overall, the fMRI evidence to date shows that cannabis use is associated with changes in partly overlapping brain pathways implicated in prominent neuroscientific theories of addiction (e.g., striatum, PFC, posterior cingulate cortex [PCC]) (7). Further, this evidence shows that different brain function in cannabis users – in striatal, orbitofrontal, amygdala, occipital, and insular pathways implicated in neuroscientific theories of addiction (7) – is associated with greater cannabis exposure and related problems, including craving, loss of control over use, grams per month, and THC metabolites (10).

To date, there continues to be substantial variability in the specific brain regions/circuits that identify altered brain activity in fMRI cannabis cue-induced craving studies. The variability likely reflects methodological differences (e.g., heterogeneity of the samples in levels of cannabis use) and limitations of studies. About half of the fMRI literature to date did not include control groups (13), only a few studies correlated brain function during cue-induced craving with metrics of cannabis exposure or cannabis-related problems (10), and many studies did not adjust analyses for potential confounders (e.g., nicotine and alcohol exposure, abstinence duration) (for a review, see Sehl and colleagues, 2021 (10)). Further, examination of fMRI cannabis cue-induced craving in participants who meet criteria for moderate-to-severe CUD, are non-treatment seeking and report having attempted to reduce or quit using cannabis (10), is relevant, since this represents the majority of cannabis consumers.

This study aimed to overcome the limitations of the literature to date. The primary aim was to comparebrain function during cannabis cue-induced craving fMRI task, for the first time in individuals meeting criteria for moderate-to-severe CUD and who self-report previous attempts to cut down or quit use, with non-using controls. In line with the emerging literature (14) and with neuroscientific theories of addiction(7), we hypothesized greater activity during cannabis vs neutral cues in the striatum, PFC (i.e., OFC, anterior cingulate cortex ACC], medial frontal gyrus [MFG]), and visual and parietal regions (i.e., precuneus, PCC) in participants within the moderate-to-severe CUD group compared to controls. Our secondary aim was to explore associations between brain activity during cue-induced craving and CUD symptom severity, subjective craving, and arousal ratings in response to cannabis cues, as well as additional metrics of cannabis exposure and mental health symptom scores, accounting for recent cannabis, alcohol, and cigarette use.

## 2 Methods and materials

The study is nested within a larger project where methods were pre-registered www.isrctn.com/ISRCTN76056942. The study was approved by the Human Research Ethics Committee at the Australian Catholic University (HREC:2019-71H).

### 2.1 Advertisement

Participants were recruited from the Melbourne metropolitan area, Australia, via public platforms (e.g., Facebook, flyers in the general community/university campuses). Advertisements for study participation included a link that directed community members to a ∼25-minute online survey to screen participants against the study’s eligibility criteria.

### 2.2 Participant selection

Participants were neurologically healthy adults screened against key inclusion and exclusion criteria. Inclusion criteria in the cannabis use group required: (i) endorsing a moderate-to-severe CUD based on the Structured Clinical Interview for DSM-5 Research Version (SCID-5-RV), (ii) daily/near-daily cannabis use over the past 12 months, and (iii) attempt to quit or cut down at least once over the past 24 months. Supplemental Data details further inclusion and exclusion criteria and eligibility processes for all participants.

### 2.3 Assessment procedure

All assessments lasted approximately 4 to 6 hours and were conducted by trained researchers and students at Monash Biomedical Imaging in Clayton, Victoria, Australia. All participants gave written informed consent prior to participation. Assessment included questionnaires administered via Qualtrics Version XM (https://www.qualtrics.com), face-to-face semi-structured interviews for substance use and mental health profiling, and cognitive testing (e.g., IQ) as detailed in Supplemental Data. The MRI scan included a structural T1-weighted image analyzed for registration purpose, and an fMRI cue-induced craving task during which participants watched cannabis images and visually matched control images. The assessment completion comprised a debrief and voucher reimbursement (i.e., Coles/Myers) of $100 for controls and $150 for the CUD group. Additional reimbursement in the CUD group was to compensate for further research activities as part of the broader study. Participants’ data collection took place between October 2019 and July 2022.

### 2.4 fMRI Cue Induced Craving Task

An event-related cue-induced craving fMRI task (Figure 1) was designed to measure brain activity while participants viewed 30 cannabis-related images depicting paraphernalia and use behaviors and 30 neutral images of stationary items or cooking utensils. Each image was displayed for 4 seconds, with a fixation cross appearing between images for an average of 4 seconds (±2), and the task lasted about 10 minutes. See Supplemental Data for additional detail on the fMRI task and participant instruction.

**Figure 1.**
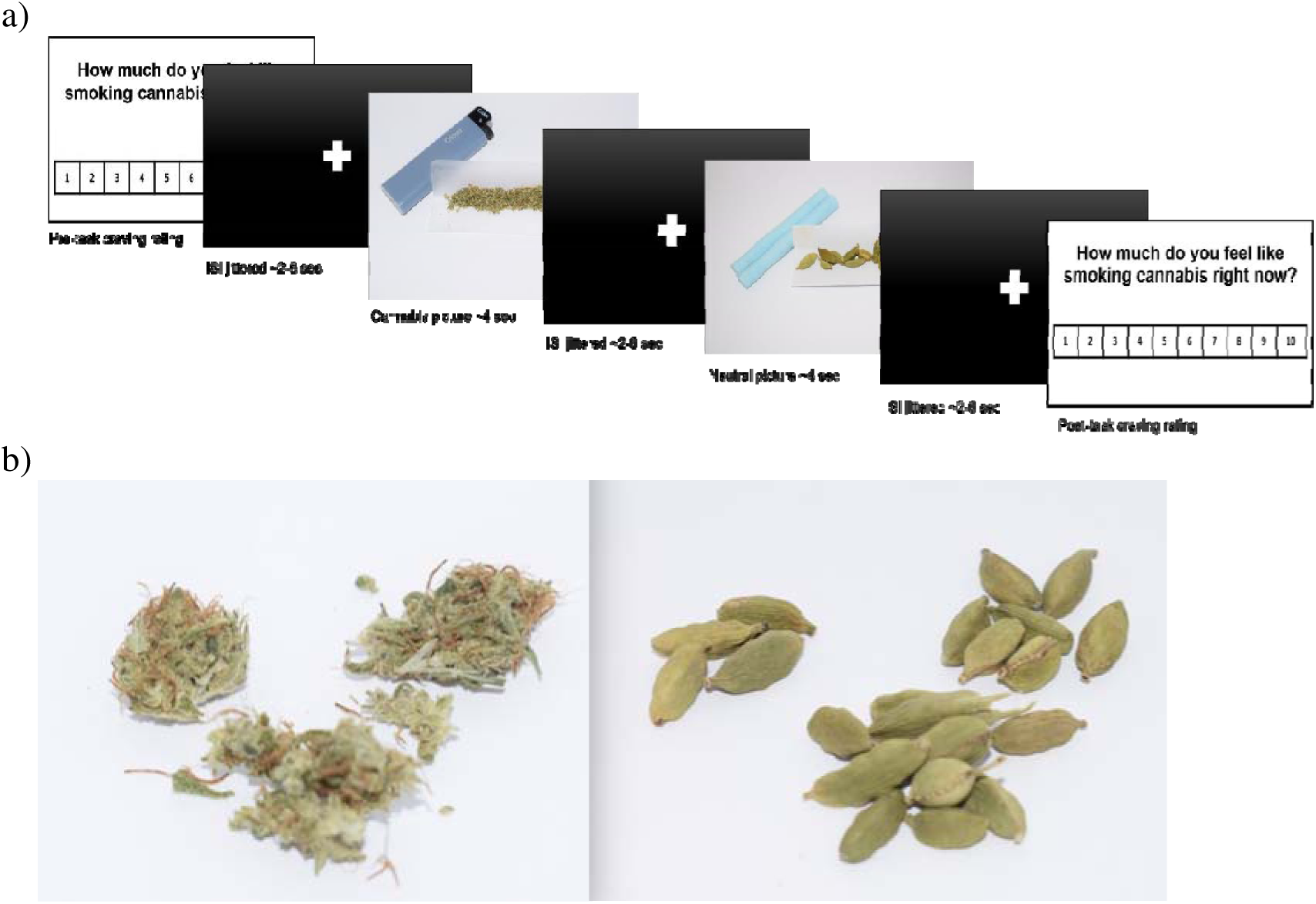
Example of fMRI Cue-Reactivity Task Parameters, including: a) Stimulus get a presentation, whereby a craving rating was presented at the start and completion of the task asking participants to rate “How much do you feel like smoking cannabis right now?” on a scale of 1-10. A total of 60 images were presented with a varying duration in seconds (sec) of inter-stimulus intervals (ISIs) shown in the black screen with a centred white ‘+’; and b) samples of a cannabis image (left) and of a neutral image.

### 2.5 Assessement of valence and craving: cannabis and neutral cues

Immediately before and after completion of the fMRI task, participants rated their arousal (0=“calm” to 10=“excited”) and affective valence (0=“unpleasant” to 10=“pleasant”) on a 10-point Likert scale and their cannabis cravings using the VAS question “how much do you feel like smoking cannabis right now?” (0=“not at all” to 10 “extremely”).

### 2.6 fMRI data analysis

First-level analyses were run on MATLAB (version R2018a) with SPM version 12 (http://fil.ion.ucl.ac.uk/spm/) using a general linear model (GLM) to quantify the relationship between the observed event-related blood-oxygen-level-dependent (BOLD) signals and two regressors (i.e. cannabis and neutral images). The six motion estimates (i.e., translation and rotation on three axes: x, y, z), their derivatives and global signal were entered as covariates of no interest. Second-level analyses in the entire sample examined task effects relate to cannabis-specific activation, using contrasts defined as cannabis > neutral, and cannabis < neutral (Figure 1, Supplemental Table 2, Supplemental Data).

Second, whole-brain group differences were examined via a GLM (i.e., cannabis > neutral and cannabis < neutral), with age and sex as covariates. A whole brain analysis was run using GLM with group as a factor (i.e., CUD vs controls). To ensure the accuracy of the results, we applied for each contrast, applying non-parametric threshold-free cluster enhancement (TFCE) (15) with 10,000 permutations, family-wise error (FWE) corrected for multiple comparisons at *p* < .05. For a significant cluster, we extracted the contrast beta values using a spherical ROI with a 5 millimeters radius centered on the peak voxel. The beta values were used to perform correlations between brain activity and behavioral data.

### 2.7 Statistical analyses

All further analyses were run using SPSS version 29. Chi-squared tests were run to compare groups by sex. Mann-Whitney *U* tests were run to compare groups for scalar and continuous variables (i.e., IQ, substance use, mental health, craving, withdrawal, mean valence and arousal ratings for cannabis and control images).

To analyze group differences in the mean affective valence and arousal ratings of images used in the fMRI cue-induced craving task, we first calculated the inter-rater reliability of all the cannabis and neutral images, respectively. We did this to determine the fit of utilizing the two groups’ mean scores of self-reported ratings of the images. Cronbach’s α>.95 indicated that ratings for all the cannabis images and all the neutral images were strongly correlated.

Therefore, using participants’ mean scores for all image ratings was appropriate to create a variable of affective valence and arousal rating of cannabis and neutral images, respectively. Group differences in mean ratings were examined using non-parametric Mann-Whitney *U* tests. Cohen’s *d* effect sizes were calculated for group differences in subjective craving and arousal/valence ratings comparisons using Psychometrica’s effect size calculator for non-parametric tests.

Spearman rank correlations were performed to examine the association between differences in brain function (i.e., β coefficients from the peak activation voxels with a kernel size 5 mm from all the significant clusters) and number of CUD symptoms, variables directly relevant for cue-induced craving (i.e., subjective craving), arousal ratings of cannabis images, and explore associations with CUD symptoms, THC-COOH:creatinine levels in urine, mental health symptom scores (i.e., Cannabis Withdrawal Scale, Beck Depression Inventory, State Anxiety Inventory, CAPE positive psychotic symptoms).

The impact of potential confounders (i.e., cigarettes used/past month, alcohol standard drinks/past month and hours since cannabis was used prior to testing) was accounted for via means of residualizing these from the β coefficients. Spearman’s rank correlations were performed with and without outliers, defined using the IQR (interquartile range) method (i.e., low outliers: Q1 – (3 * IQR); high outliers: Q3 + (3 * IQR) (Supplemental Data; sTables 3-4).

## 3 Results

Table 1 overviews the sample characteristics. The overall sample included 108 participants (35 females and 73 males) with a median age of 25 (range: 18-56 years). Of these, 65 participants were in the moderate-to-severe CUD group and 43 were controls.

**Table 1.**
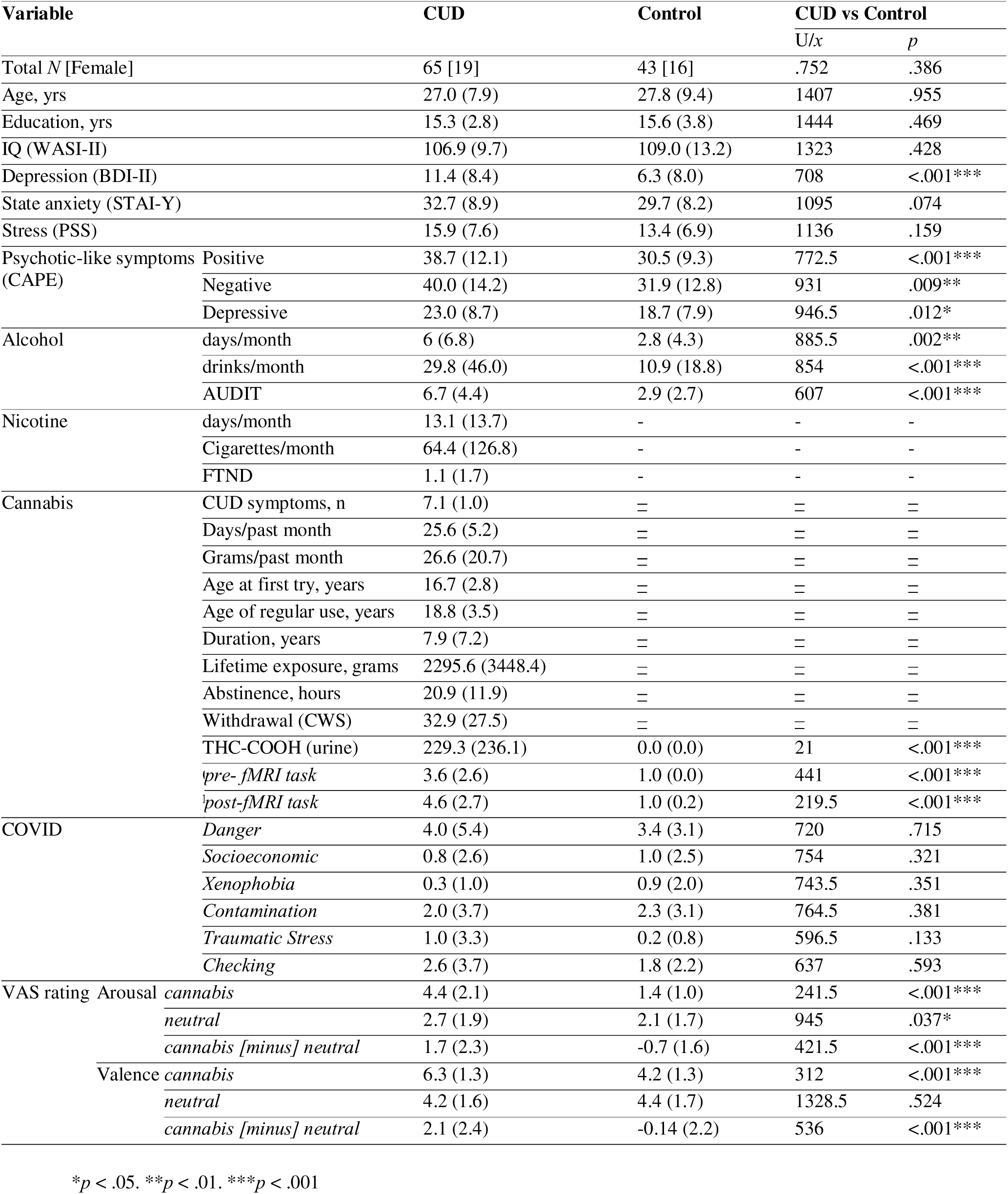
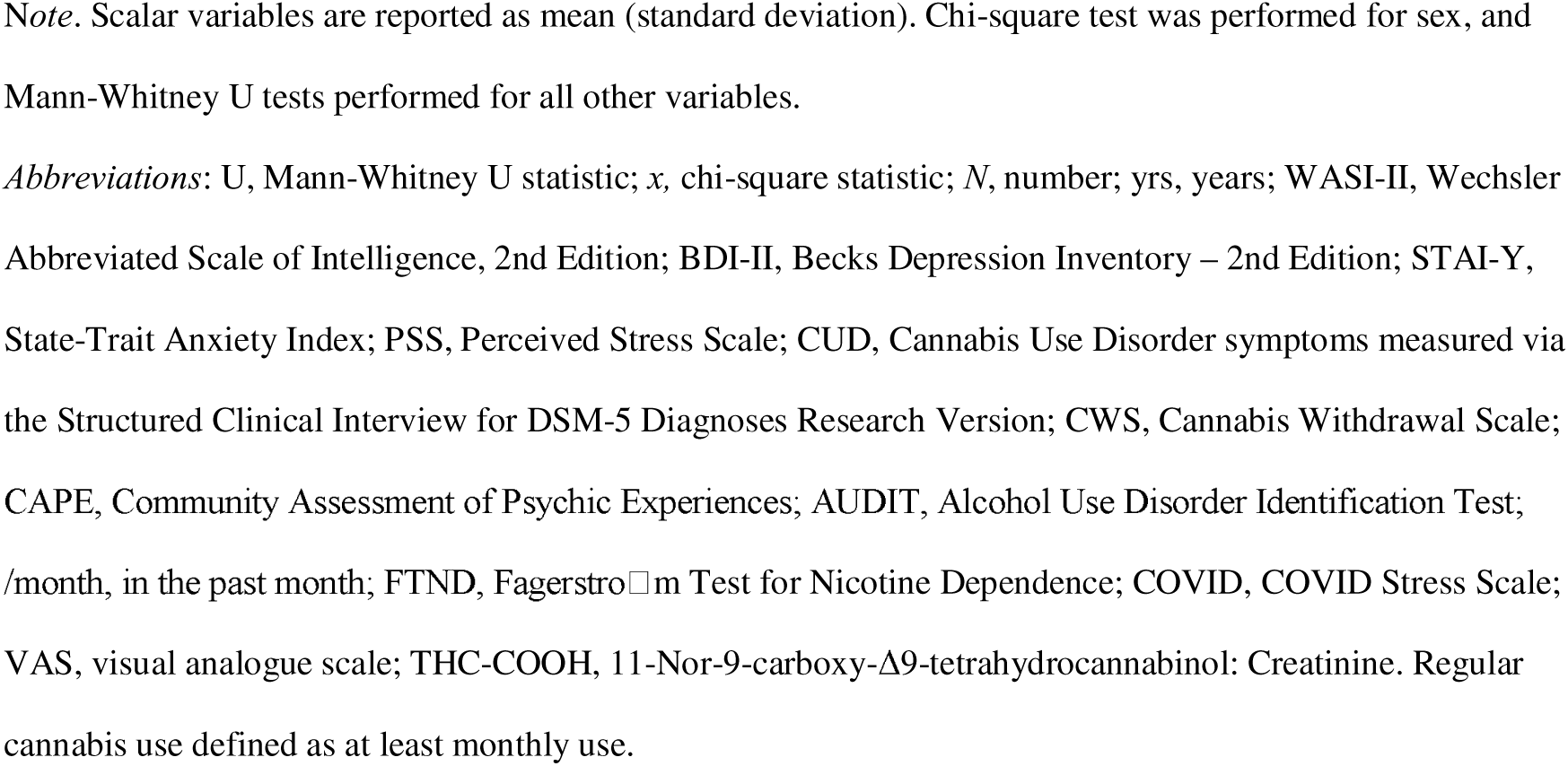
Sample Characteristics: Socio-demographic Data, Education, IQ, Mental Health, Alcohol and Nicotine Use and Dependence Levels

| Variable |  | CUD | Control | CUD vs Control |  |
| --- | --- | --- | --- | --- | --- |
|  |  |  |  | U/x | p |
| Total N [Female] |  | 65 [19] | 43 [16] | .752 | .386 |
| Age, yrs |  | 27.0 (7.9) | 27.8 (9.4) | 1407 | .955 |
| Education, yrs |  | 15.3 (2.8) | 15.6 (3.8) | 1444 | .469 |
| IQ (WASI-II) |  | 106.9 (9.7) | 109.0 (13.2) | 1323 | .428 |
| Depression (BDI-II) |  | 11.4 (8.4) | 6.3 (8.0) | 708 | <.001*** |
| State anxiety (STAI-Y) |  | 32.7 (8.9) | 29.7 (8.2) | 1095 | .074 |
| Stress (PSS) |  | 15.9 (7.6) | 13.4 (6.9) | 1136 | .159 |
| Psychotic-like symptoms (CAPE) | Positive | 38.7 (12.1) | 30.5 (9.3) | 772.5 | <.001*** |
|  | Negative | 40.0 (14.2) | 31.9 (12.8) | 931 | .009** |
|  | Depressive | 23.0 (8.7) | 18.7 (7.9) | 946.5 | .012* |
| Alcohol | days/month | 6 (6.8) | 2.8 (4.3) | 885.5 | .002** |
|  | drinks/month | 29.8 (46.0) | 10.9 (18.8) | 854 | <.001*** |
|  | AUDIT | 6.7 (4.4) | 2.9 (2.7) | 607 | <.001*** |
| Nicotine | days/month | 13.1 (13.7) | - | - | - |
|  | Cigarettes/month | 64.4 (126.8) | - | - | - |
|  | FTND | 1.1 (1.7) | - | - | - |
| Cannabis | CUD symptoms, n | 7.1 (1.0) | = | = | = |
|  | Days/past month | 25.6 (5.2) | = | = | = |
|  | Grams/past month | 26.6 (20.7) | = | = | = |
|  | Age at first try, years | 16.7 (2.8) | = | = | = |
|  | Age of regular use, years | 18.8 (3.5) | = | = | = |
|  | Duration, years | 7.9 (7.2) | = | = | = |
|  | Lifetime exposure, grams | 2295.6 (3448.4) | = | = | = |
|  | Abstinence, hours | 20.9 (11.9) | = | = | = |
|  | Withdrawal (CWS) | 32.9 (27.5) | = | = | = |
|  | THC-COOH (urine) | 229.3 (236.1) | 0.0 (0.0) | 21 | <.001*** |
|  | *pre-fMRI task* | 3.6 (2.6) | 1.0 (0.0) | 441 | <.001*** |
|  | *post-fMRI task* | 4.6 (2.7) | 1.0 (0.2) | 219.5 | <.001*** |
| COVID | *Danger* | 4.0 (5.4) | 3.4 (3.1) | 720 | .715 |
|  | *Socioeconomic* | 0.8 (2.6) | 1.0 (2.5) | 754 | .321 |
|  | *Xenophobia* | 0.3 (1.0) | 0.9 (2.0) | 743.5 | .351 |
|  | *Contamination* | 2.0 (3.7) | 2.3 (3.1) | 764.5 | .381 |
|  | *Traumatic Stress* | 1.0 (3.3) | 0.2 (0.8) | 596.5 | .133 |
|  | *Checking* | 2.6 (3.7) | 1.8 (2.2) | 637 | .593 |
| VAS rating | Arousal *cannabis* | 4.4 (2.1) | 1.4 (1.0) | 241.5 | <.001*** |
|  | *neutral* | 2.7 (1.9) | 2.1 (1.7) | 945 | .037* |
|  | *cannabis [minus] neutral* | 1.7 (2.3) | -0.7 (1.6) | 421.5 | <.001*** |
|  | Valence *cannabis* | 6.3 (1.3) | 4.2 (1.3) | 312 | <.001*** |
|  | *neutral* | 4.2 (1.6) | 4.4 (1.7) | 1328.5 | .524 |
|  | *cannabis [minus] neutral* | 2.1 (2.4) | -0.14 (2.2) | 536 | <.001*** |

\* $p < .05$ . \*\* $p < .01$ . \*\*\* $p < .001$
*Note.* Scalar variables are reported as mean (standard deviation). Chi-square test was performed for sex, and Mann-Whitney U tests performed for all other variables.

### 3.1 Sample socio-demographic, mental health and alcohol/nicotine use characteristics

Groups did not significantly differ in sex, age, IQ, state anxiety, perceived stress and COVID-19 related stress. Compared to controls, people with a CUD had significantly higher depressive symptoms, positive, negative, and depressive psychotic-like symptoms, and greater past month alcohol exposure. Alcohol-related problems (AUDIT) were significantly higher in the CUD group. Nicotine dependence (FTND score ≥ 3) was endorsed by 10 participants with a CUD, while no control participants reported using nicotine.

### 3.2 Cannabis consumption and related problems

Participants in the CUD group endorsed a median of 7 CUD symptoms (range 4-to-11), with 73.4% of them experiencing a severe CUD and the remainder a moderate CUD. Mean cannabis consumption in the CUD group was 26.6 grams over 30 days in the past month – approximately 0.7 grams daily. Mean cumulative lifetime use was 898 grams, calculated as: the mean age of first cannabis use was 16 years, at least monthly use commencing at 18 years, and a mean duration of regular use of 7.9 years.

Participants abstained from cannabis for a mean of 20.9 hours before testing. Cannabis withdrawal symptoms on the CWS were a mean of 32.9 of a possible score of 190, and ranged from no withdrawal to moderate withdrawal. Corroborating self-reported cannabis use, levels of THC-COOH:creatinine in urine were detected among the CUD group but not in any controls.

### 3.3 Subjective craving pre-to-post cue-induced craving fMRI task and arousal/valence ratings of images

Subjective craving examined pre and post the cue-induced craving fMRI task was significantly higher in participants with a CUD compared to controls (*d = 1.39, p < .001 and d = 2.00, p < .001*, respectively). In addition, the CUD group exhibited a greater increase between time points (*d* = *0.91*, *p* < *.001*).

People with a CUD compared to controls rated *cannabis images* as triggering greater arousal levels, and as having positive valence instead of neutral valence, with strong effect sizes (*d = 1.88* and *d = 1.65*, respectively). For *neutral images*, arousal ratings were significantly higher in the CUD compared to the control group, with a small effect size (*d = 0.41*). Neutral image valence ratings did not differ significantly between groups. People with a CUD compared to controls had significantly greater arousal and valence ratings of *cannabis (minus) neutral images* (*d*=*1.35*, *p<.001* and *d* =*1.10, p*<.*001*, respectively).

### 3.4 Group differences in brain activity during the cannabis cue-induced craving fMRI task

Compared to controls, participants with a moderate-to-severe CUD showed greater BOLD activity in a range of areas (*p<.05*; FWE corrected) while viewing cannabis vs neutral images (Figure 2 and Table 2). Greater activity emerged in visual/attentional areas (i.e., superior/middle occipital and fusiform gyri, and calcarine sulcus); in PFC regions (i.e., medial OFC, ACC), temporal regions (i.e., hippocampus, middle temporal gyrus), as well as posterior cingulate, parietal cortices (e.g., inferior parietal, postcentral and supramarginal gyri) and cerebellum.

**Figure 2.**
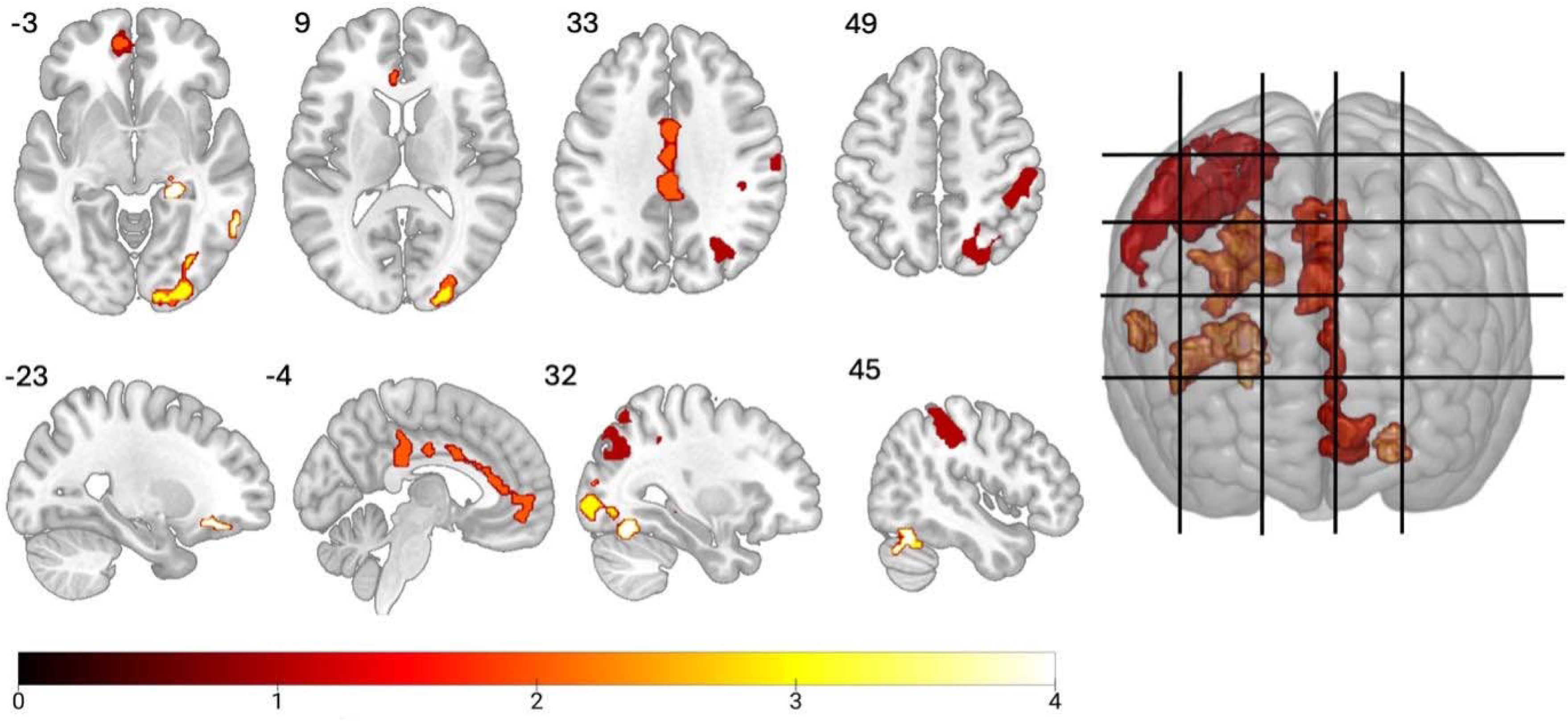
Overview of significantly greater brain activity (*p < .05*; FWE-corrected in axial, sagittal and 3D views) in participants with a moderate-to-severe CUD compared to controls while viewing cannabis>neutral pictures. Color bar represents the *t* values for Threshold Free Cluster Enhancement (TFCE). These functional maps are overlaid on an MNI space template image. Gridlines represent the displayed slices.

**Table 2.**
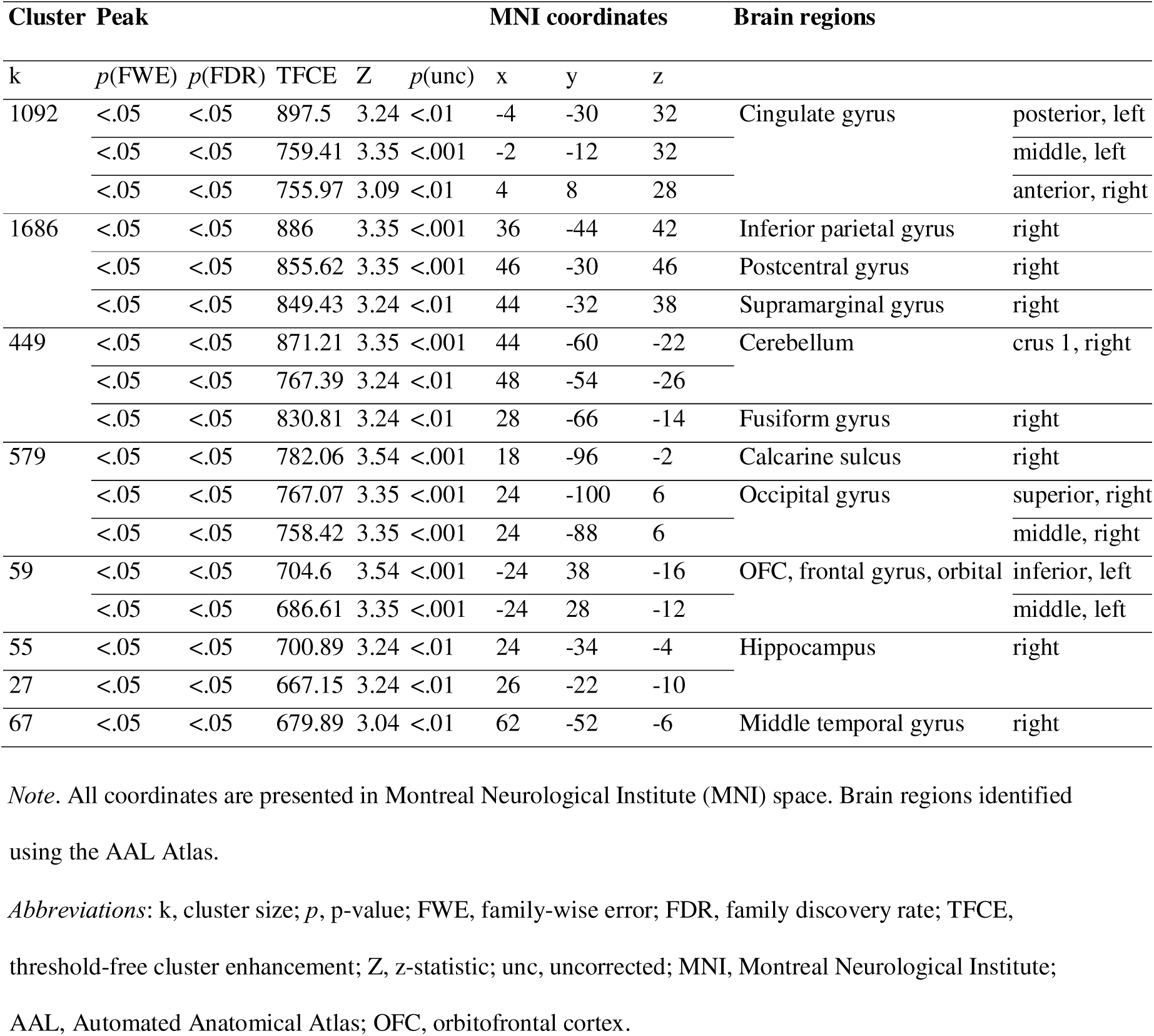
Group differences in neural cannabis cue-induced craving fMRI task (cannabis **> neutral images, *p < .05*, FWE corrected)**

### 3.5 Brain-behavior correlations

Table 3 shows correlations between brain activity and behavioral variables, accounting for cigarettes used/past month, alcohol standard drinks/past month and hours since cannabis was last used prior to testing. Within the CUD group, there were positive correlations between greater superior occipital cortex activity and arousal ratings of cannabis (*minus*) neutral images (VAS scores; rho = 0.33, *p < .05*), and withdrawal (CWS scores; rho = 0.32, *p < .05*). THC-COOH:creatinine levels negatively correlated with activity of the ACC and inferior parietal cortex (rho = -0.28, *p < .05* and rho = -0.30, *p < .05*, respectively). These correlations were still significant after outlier removal. Additional correlations did not survive outlier removal or were non-significant.

**Table 3.**
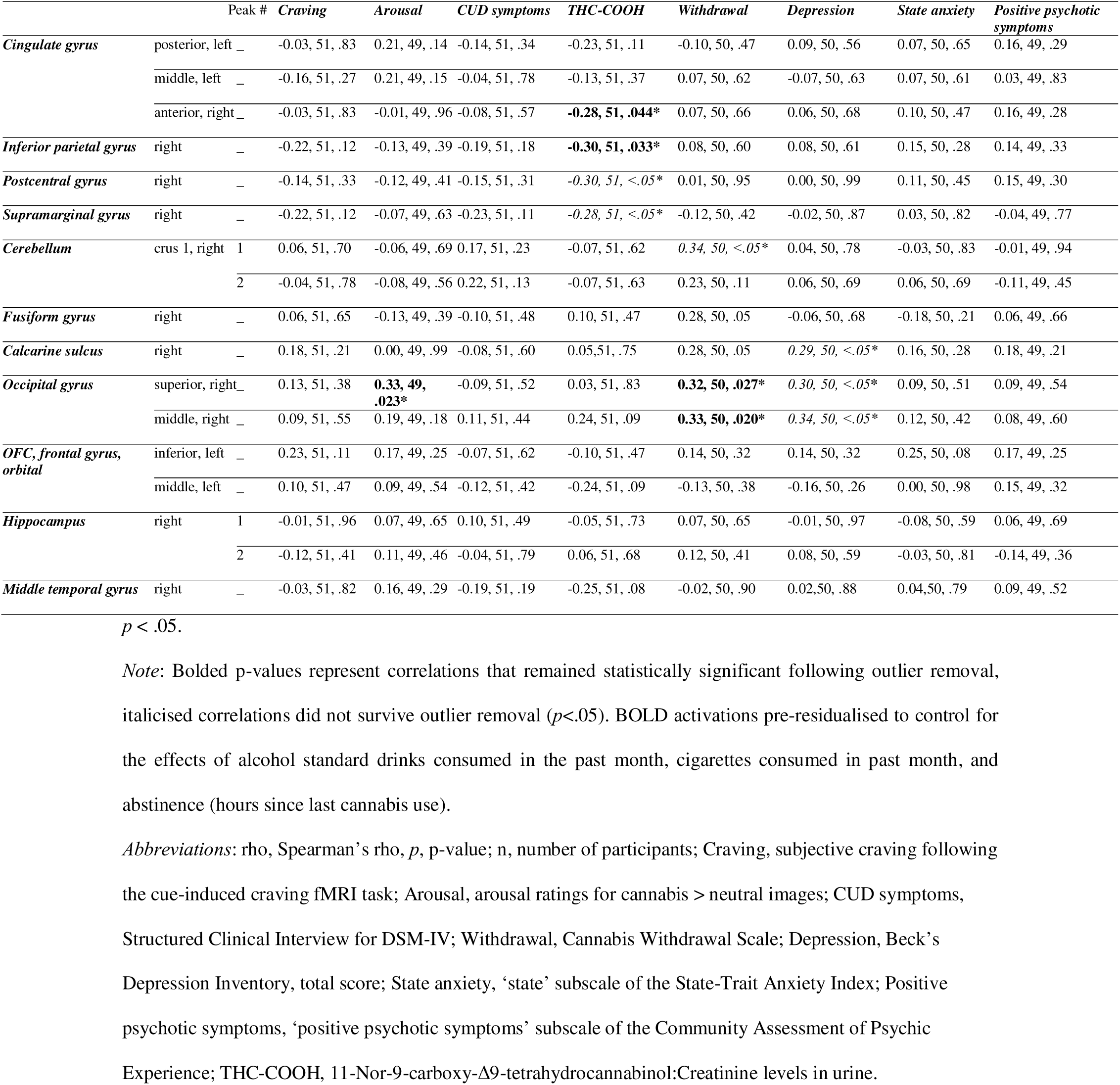
Brain-behaviour correlations in participants with a CUD (rho, n, *p*).

## 4 Discussion

This is the first and largest fMRI study to our knowledge, to examine brain function during cue-induced craving in people with moderate-to-severe CUD who tried to cut down or quit and comprehensively characterized for cannabis use levels, in comparison to controls. We found that compared to controls, during cue-induced craving CUD participants exhibited greater activity in the ACC and inferior parietal cortex, in correlation with urinary THC-COOH levels. The CUD group also showed greater brain activity in the visual regions, which correlated with greater arousal ratings of cannabis images, and withdrawal scores. Further, participants with a CUD, compared to controls, showed greater activity in the OFC, temporal regions (i.e., hippocampus, middle temporal gyrus), posterior cingulate, parietal cortices (e.g., inferior parietal, postcentral and supramarginal gyri) and cerebellum.

Compared to controls, people with CUD exhibited greater PFC activity during cue-induced craving, encompassing the OFC and the ACC, both regions implicated in regulating responses to salient drug related stimuli (12). This finding was consistent with our hypothesis, fMRI evidence on cue-induced craving tasks on people who use cannabis (14), other substances (16), and neuroscientific theories of addiction (7). The OFC regulates limbic-striatal regions implicated in reward processing, motivation and effortful control (12). Thus, greater OFC cue-reactivity may reflect an increased attribution of salience and reward expectancy to cannabis cues (17). Meanwhile, the ACC is implicated in supporting awareness of negative emotional states (7). Therefore, the correlation between greater ACC activity and lower THC metabolites in urine might indicate that participants whose circulating levels of THC are lower might experience greater reactivity to cue-induced craving, possibly predating a need for intaking more THC.

The most robust aspect of cue-induced craving (to date) is arguably visual processing due to the visual exposure to cues. Indeed we found that people with a CUD compared to controls, showed stronger activity in occipital areas implicated in the processing of higher-order visual stimuli (18); in line with previous work in people who use cannabis (10) and other substances (e.g., nicotine, alcohol) (19). Of note, occipital activity significantly correlated with greater arousal to cannabis images, and withdrawal. Importantly, post-hoc analyses confirmed that arousal and withdrawal were not colinear and both independently contributed to explaining the variance of the OFC activity (i.e., they were not co-linear). As occipital regions mediate internally directed attention (20), occipital reactivity may reflect greater attention to people’s own reactions to cannabis cues (21). This notion needs to be tested using objective measures of attention to cannabis cues (e.g., eye tracker), interoception and correlating such metrics with visual cortical activity (22).

We also found greater cerebellar activity in people with a CUD while viewing cannabis versus neutral images, in line with previous evidence in people who use cannabis (13), and other substances (e.g., alcohol (23)). Notably, the cerebellum is a core component of the addiction neurocircuitry (7) and exhibits a high density of cannabinoid type 1 receptors, to which THC, the main psychoactive compound of cannabis, binds to exert psychoactive effects (24). The cerebellum is also implicated in drug-associated cue-memory and cue-elicited cerebellar activity may reflect an anticipatory response to cannabis cues (24).

The greater activation of the hippocampus and medial temporal areas in CUD participants compared to controls is in line with fMRI evidence on cue-induced craving in people who use cannabis (10). Hippocampal and temporal regions are implicated in stress and verbal/visuospatial learning and memory processes (25) known to be altered in this population (26). Thus, cue-induced craving might have triggered greater activation of stress and cue-related memory circuits (25).

The activity of parietal regions was also greater in the CUD group compared to controls (i.e., inferior parietal, postcentral, supramarginal gyri). The reported alteration in parietal activity is consistent with fMRI cue-induced craving evidence in people who use cannabis and other substances (10, 27, 28) and prominent neuroscientific theories of addiction (7). Interestingly, the postcentral gyrus, reportedly enhanced in cannabis users, is an integral part of the sensorimotor network implicated in the integration of sensory stimuli with emotions and memories (29, 30) while the supramarginal gyrus is implicated in episodic memory (31), also altered in people who use cannabis (32). Therefore, exposure to cannabis cues in CUD may aid recollection of previous cannabis use experiences and their context (e.g., associated emotions, sensory experiences, time, place) and trigger craving and disinhibition of cannabis use behavior.

In contrast with the hypothesis, there was no group difference in brain activity in the striatum, a key hub for reward processing in the addiction neurocircuitry (7). This partially aligns with previous drug cue-induced craving fMRI tasks, where both group differences and lack thereof have been reported (13). The function of the striatum is difficult to measure with neuroimaging modalities/techniques due to its high concentration of iron (33). It is possible that region-of-interest data acquisition and analysis approaches (instead of the whole-brain exploratory approaches used in this work) are required to measure with precision changes in brain function of the striatum. Additionally, future work is required to determine if and how other variables are associated with brain activity in CUD (e.g., age, sex, cannabis use patterns such as age of onset).

Subjective craving significantly increased pre-to-post the cue-induced craving fMRI task, and the CUD group (not controls) reported greater subjective valence/arousal in relation to cannabis versus neutral images, suggesting that the stimuli and the cue-exposure task might have triggered increases in craving. Yet, subjective craving did not correlate with brain function, consistent with previous evidence (13). However, the literature to date has reported both significant and non-significant correlations between craving and cue-induced craving-related brain function (13). The relationship between cue-elicited brain function and subjective craving may be modulated by other factors e.g., perceived availability of cannabis use post-presentation of cannabis cues (34), and varying levels of CUD symptoms, which were unmeasured. It is possible that inter-individual variability in craving was too low to capture its association with brain function (i.e., a narrow range of subjective craving), and meta-regressions of the literature may be needed to explore this association.

Overall, the regions involved in cue-induced craving in CUD are critical components of three distinct systems previously implicated in craving (35): the visual system to process the cannabis cues (e.g., occipital regions), the motivational/inhibitory systems implicated in top-down control (e.g., OFC, ACC and parietal regions), and the subcortical temporo-limbic system implicated in stress and cue-related memory activation (e.g., hippocampus, cerebellum). Functional connectivity analyses are required to examine the functional communication between these systems.

The findings from this study need to be considered in light of several methodological limitations. First, the cross-sectional design prevents understanding of whether group differences predated CUD (36), or changed over time as people progress to greater or lower CUD severity, relapse or following prolonged abstinence. Longitudinal studies are required to test this notion. Second, we lacked objective proxies of craving (e.g., skin conductance) and of stress (e.g., cortisol); which should be integrated in future studies to precisely map the multimodal mechanisms of cue-induced craving in CUD. Since the study was restricted to people with moderate-to-severe CUD and with a history of attempting to quit or cut down due to a focus on cannabis users who experience difficulties with use, the findings may not generalize to other groups of cannabis users (e.g., mild CUD or no CUD).

Fourth, the cue-induced craving fMRI task used herein robustly induced craving and triggered cue-reactivity (37) and was robust in that cannabis and neutral stimuli were well matched according to various characteristics (e.g., complexity, size, brightness, color, resolution) (38). However, specific/personalized cues might be required to trigger cue-induced craving for a more naturalistic design that generalize to individual experiences outside the scanner (39). Finally, the study lacked control images with rewarding properties (e.g., monetary) that also recruit brain reward pathways altered in people who use cannabis (40). The findings need to be replicated using additional rewarding stimuli, to disambiguate whether cue-induced craving was specific to cannabis images or applies to rewarding images generally.

Reactivity to neural cues has been associated with relapse in other substance use disorders Our findings provide a preliminary foundation for informing treatment targets for individuals with more severe CUDs who are vulnerable to relapse (42). Brain-based interventions that target brain functional alterations specific to the individual during cue-induced craving - such as fMRI-based neurofeedback, and transcranial magnetic stimulation (43) - could prove useful to reduce individually-specific reactivity to cannabis cues.

Therefore, such treatments could inform the development of personalized interventions to prevent relapse and support long-term recovery in individuals who experience CUDs. Such brain-based interventions could be combined with therapies that strengthen ‘top-down’ executive control or reduce ‘bottom-up’ automatic responses to cues (e.g., cognitive bias modification (44), mindfulness-based relapse prevention (45)). Our findings also support the investigation of interventions that subjectively devalue cannabis by reducing its expected reward whilst increasing the relative valuations of non-drug rewards.

In conclusion, the findings suggest that cannabis cue-induced craving in moderate-to-severe CUD who tried to cut down or quit, is associated with greater activity of the prefrontal, temporal, and parietal regions of the addiction neurocircuitry (12). It is also associated with additional activity in the occipital regions implicated in higher-order visual/attention processing, and with cannabis withdrawal, depressive symptom scores, and arousal elicited by the images administered during the cue-induced craving fMRI task. The neurobiology of cannabis cue-induced craving in CUD may affect fronto-temporo-parietal neurocircuitry overlapping with other SUDs (i.e., alcohol, nicotine, opioids, cocaine; (41)), and may (in part) align with prominent neuroscientific theories of addiction (12). Our findings have implications for the development of psychological and brain-based interventions that target the function of these regions to decrease craving, cannabis dosage and the severity of CUD.

## Data Availability

The datasets generated during the current study will be available upon reasonable request from Dr Valentina Lorenzetti, Principal Investigator. Data will include relevant group allocations and outcome variables and will be anonymised. Data will be available either as it is published, or on request. A time limit will not be set on the duration of availability. Data will be shared with anyone who wishes to access it, for meta-analyses or other pre-approved purposes, via email. All participants provided informed consent. All data is de-identified.

## Acknowledgments

We thank all participants for contributing to the project with their data and time. We acknowledge Ms Natalie DeBono, Dr Leonie Duehlmeyer and Dr Penny Hartman for contributing to the management of the setting up of the project. We acknowledge Ms Stephanie Antopolous, Ms Claire Chua, Dr Leonie Duehlmeyer, Mr Lachlan Grant, Ms Kirsty Kearney, Dr Magdalena Kowalczyk, Ms Emily Robinson, Ms Elizabeth Sharp, and Ms Danielle Tichelaar, for their contribution to data collection. We acknowledge Dr Chandni Hindocha for her contribution to taking pictures of cannabis products, which were used in the fMRI cue-induced craving task. We acknowledge Professor Shanlin Fu and the team at the Drugs and Toxicology Group, Centre for Forensic Science, University of Technology Sydney, for conducting urine toxicology analyses. We also acknowledge Ms Anastasia Paloubis who provided assistance in the formatting of the manuscript and in the operations required to submit the work.

## Acknowledgments

Valentina Lorenzetti was supported by an Al and Val Rosenstrauss Research Fellowship (2022–2026), and by a National Health & Medical Research Council (NHMRC) Investigator Grant (2023-2027, ID 2016833) and an Australian Catholic University competitive scheme. The work within the Neuroscience of Addition and Mental Health Program, Healthy Brain and Mind Research Centre was supported via an ACU competitive scheme. Emillie Beyer, Hannah Sehl, Hannah Thomson and Marianna Quinones Valera were funded by Australian Government Research Training Program (RTP) Stipend scholarships. Victoria Manning has received funding from the National Health and Medical Research Council (NHMRC), VicHealth, the Department of Health Victoria, the Victorian Responsible Gambling Foundation, the Victorian Medical Research Acceleration Fund, NSW Health, the National Centre for Clinical Research on Emerging Drugs (NCCRED), HCF, and philanthropic organizations.

## Previous presentation

Valentina Lorenzetti, presented these results on the 29 of May 2024, at the Department of Psychiatry, The University of Melbourne

## Authors contribution

- All authors edited the manuscript.
- AC managed all the operations of the study and edited the manuscript.
- AHA drove all aspects of the fMRI analyses and QCs, and created figures with fMRI results, with general direction on the technical aspects from CS, GP and on the theoretical aspects from VL.
- CS provided high-level and ongoing input on all aspects of the study with a focus on the neuroimaging technical aspects.
- EM drove all the statistical analyses of the behavioral data, correlations, tables and figures under the general direction of VL, provided input in theoretical aspects of the manuscript and edited the manuscript.
- GP provided high-level and ongoing input on the design and conduct of the study, edited the first full draft of the manuscript and subsequent drafts, and provided high-level input into the fMRI analysis.
- HS supported the study setup. Under the PhD supervision of VL, VM and GP, HS in the context of preliminary analyses on a portion of the sample reported herein: developed the theoretical framework of the manuscript, conducted fMRI quality checks in a subsample, drove statistical analyses of the behavioral data in a subsample, and wrote an earlier version of the manuscript.
- AG, DT, EB, HS, HT, MQ supported data collection for a substantial portion of the sample.
- JC provided the fMRI cue induced craving task.
- IL, PR and GT supported the setup of the study protocol.
- LG provided high-level and ongoing input on all aspects of the study.
- VL designed and led the study as CI, supervised all students and staff involved, created the first draft of the manuscript and led all revisions.
- VM provided high-level and ongoing input on the design and conduct of the study, edited the first full draft of the manuscript and subsequent drafts, and provided high-level input into the theoretical framework and clinical relevance.

## Abbreviations

AUDIT: Alcohol use disorder identification test
BDI-II: Beck depression Inventory, version II
BOLD: Blood-oxygen level dependent
CAPE: Community assessment of psychiatric experiences
CUD: Cannabis use disorder
CWS: Cannabis withdrawal scale
DSM-IV: Diagnostic and statistical manual of mental disorders version 4
DSM-5: Diagnostic and statistical manual of mental disorders version 5
DVARS: Derivatives of root mean square variance over voxels
FD: Framewise displacement
FWE: Family-wise error
fMRI: functional magnetic resonance imaging
FTND: Fagerström test for nicotine dependence
GLM: General linear model
IQ: Intelligence quotient
MNI: Montreal Neurological Institute
MRI: Magnetic resonance imaging
OFC: Orbitofrontal cortex
PCC: Posterior cingulate cortex
PFC: Prefrontal cortex
PSS: Perceived stress scale
ROI: Region of interest
SCID-5: Structured Clinical Interview for DSM-5 Research Version
SUD: Substance use disorder
THC: Δ-9-tetrahydrocannabinol
THC-COOH: Delta-8 and/or delta-9 carboxy tetrahydrocannabinol
TFCE: Threshold-free cluster enhancement
VAS: Visual analogue scale
WASI-II: Wechsler abbreviated scale of intelligence

