## Supplemental Table 1 for "The neurocircuitry of cue-induced cannabis craving in Cannabis Use Disorder: A functional neuroimaging study"

**Supplemental Table 1. Overview of participants’ occasional exposure to illicit substances within 4 weeks from testing**

| **Substance** | **Group** | **Age** | **Sex** | **Hours before testing** | **Dosage** |
| --- | --- | --- | --- | --- | --- |
| Amyl Nitrate | Control | 20s | male | 48 | 1 dose |
| Cocaine | CUD | 20s | male | 624 | 1 gram |
|  | CUD | 30s | female | 552 | 1 gram |
|  | CUD | 20s | male | 38 | 2.5 gram |
|  | CUD | 30s | male | 392 | 0.1 gram |
|  | CUD | 20s | male | 432 | 0.3 gram |
|  | CUD | 20s | male | 229 | 0.8 gram |
|  | CUD | 20s | male | 139 | 50 mg |
| Dexamphetamine | CUD | 30s | male | 29 | 20 mg |
| Ketamine | CUD | 20s | male | 240 | 0.5 gram |
| MDMA | CUD | 30s | male | 108 | 1 dose |
|  | CUD | 20s | male | 368 | 0.5 gram |
|  | Control | 20s | male | 60 | 0.1 gram |
|  | CUD | 20s | female | 16 | 0.15 gram |
|  | CUD | 20s | male | 546 | 0.1 gram |
| Mescaline | CUD | 20s | male | 648 | 1 dose |
| Modafinil | Control | 20s | male | 432 | 50 mg |
|  | CUD | 20s | male | 96 | 0.2 gram |
| Mushrooms | CUD | 20s | male | 638 | 2 gram |
|  | CUD | 20s | male | 148 | 1.8 gram |
|  | CUD | 20s | male | 648 | 2 gram |
|  | CUD | 20s | male | 504 | 1 gram |
|  | CUD | 20s | male | 137 | 1 gram |
| Valium | CUD | 20s | female | 40 | 5 mg |
| Vyvanse | CUD | 20s | female | 28 | 45 mg |
| Xanax | CUD | 20s | female | 360 | 1 mg |
|  | CUD | 20s | male | 159 | _ |
