## Supplemental Table 2 for "The neurocircuitry of cue-induced cannabis craving in Cannabis Use Disorder: A functional neuroimaging study"

**Supplemental** **Table 2. Brain regions implicated in the fMRI cue-induced craving task across the whole sample (*p*<0.001, FWE corrected)**

| **Cluster** | **Peak** | | | | | **MNI coordinates** | | | **AAL labels** | |
| --- | --- | --- | --- | --- | --- | --- | --- | --- | --- | --- |
| **k** | **p(FWE)** | **p(FDR)** | **TFCE** | **Z** | **p(unc)** | **x** | **y** | **z** |  | |
| **cannabis > neutral pictures** | | | | | | | | | | |
| 4226 | <.001 | <.01 | 14873.95 | 3.35 | <.001 | -8 | -52 | 28 | Precuneus | left |
|  | <.001 | <.01 | 13874.41 | 3.35 | <.001 | 6 | -52 | 26 |  | right |
|  | <.001 | <.01 | 12723.94 | 3.35 | <.001 | 0 | -60 | 34 |  | left |
| 5602 | <.001 | <.01 | 13552.96 | 3.35 | <.001 | -6 | 52 | -8 | Medial OFC, left | |
|  | <.001 | <.01 | 13351.92 | 3.35 | <.001 | -4 | 42 | -2 | ACC, left | |
|  | <.001 | <.01 | 12874.29 | 3.35 | <.001 | -2 | 36 | -8 |  |  |
| 377 | <.01 | <.01 | 6584.78 | 3.35 | <.001 | -26 | -34 | -6 | Hippocampus, left | |
|  | <.01 | <.01 | 6441.6 | 3.35 | <.001 | -24 | -22 | -16 |  |  |
|  | <.01 | <.01 | 5673.34 | 3.35 | <.001 | -20 | -38 | 2 |  |  |
| 135 | <.01 | <.01 | 5996.43 | 3.35 | <.001 | 62 | -16 | 32 | Postcentral gyrus, right | |
| 37 | <.01 | <.01 | 5190.96 | 3.35 | <.001 | -30 | 6 | -16 | Amygdala, left | |
|  | <.01 | <.01 | 5124.96 | 3.35 | <.001 | -34 | 14 | -14 | Insula cortex, left | |
| 19 | <.01 | <.01 | 5168.55 | 3.35 | <.001 | -42 | 10 | -6 |  |  |
| 6 | <.01 | <.01 | 5078.22 | 3.35 | <.001 | 28 | -22 | -14 | Hippocampus, right | |
| 10 | <.01 | <.01 | 5066.02 | 3.35 | <.001 | -64 | -8 | -14 | Middle temporal gyrus, left | |
| **neutral > cannabis pictures** | | | | | | | | | | |
| 748 | <.001 | <.01 | 2426.83 | 3.35 | <.001 | 14 | -74 | 0 | Lingual gyrus, right | |
|  | <.001 | <.01 | 2387.86 | 3.35 | <.001 | 8 | -82 | 0 |  |  |
|  | <.001 | <.01 | 2315.09 | 3.35 | <.001 | 12 | -82 | 10 | Calcarine sulcus, right | |
| 149 | <.001 | <.01 | 1640.99 | 3.35 | <.001 | -38 | 6 | 30 | Inferior frontal gyrus, operculum, left | |
|  | <.01 | <.01 | 1461.85 | 3.35 | <.001 | -46 | 12 | 30 |  |  |
| 77 | <.01 | <.01 | 1459.22 | 3.35 | <.001 | -52 | 30 | 20 |  |  |
|  | <.01 | <.01 | 1430.49 | 3.35 | <.001 | -44 | 20 | 26 | Inferior frontal gyrus, triangularis, left | |
