## Supplementary figures and images for "The neurocircuitry of cue-induced cannabis craving in Cannabis Use Disorder: A functional neuroimaging study"

### Supplemental Figure 1

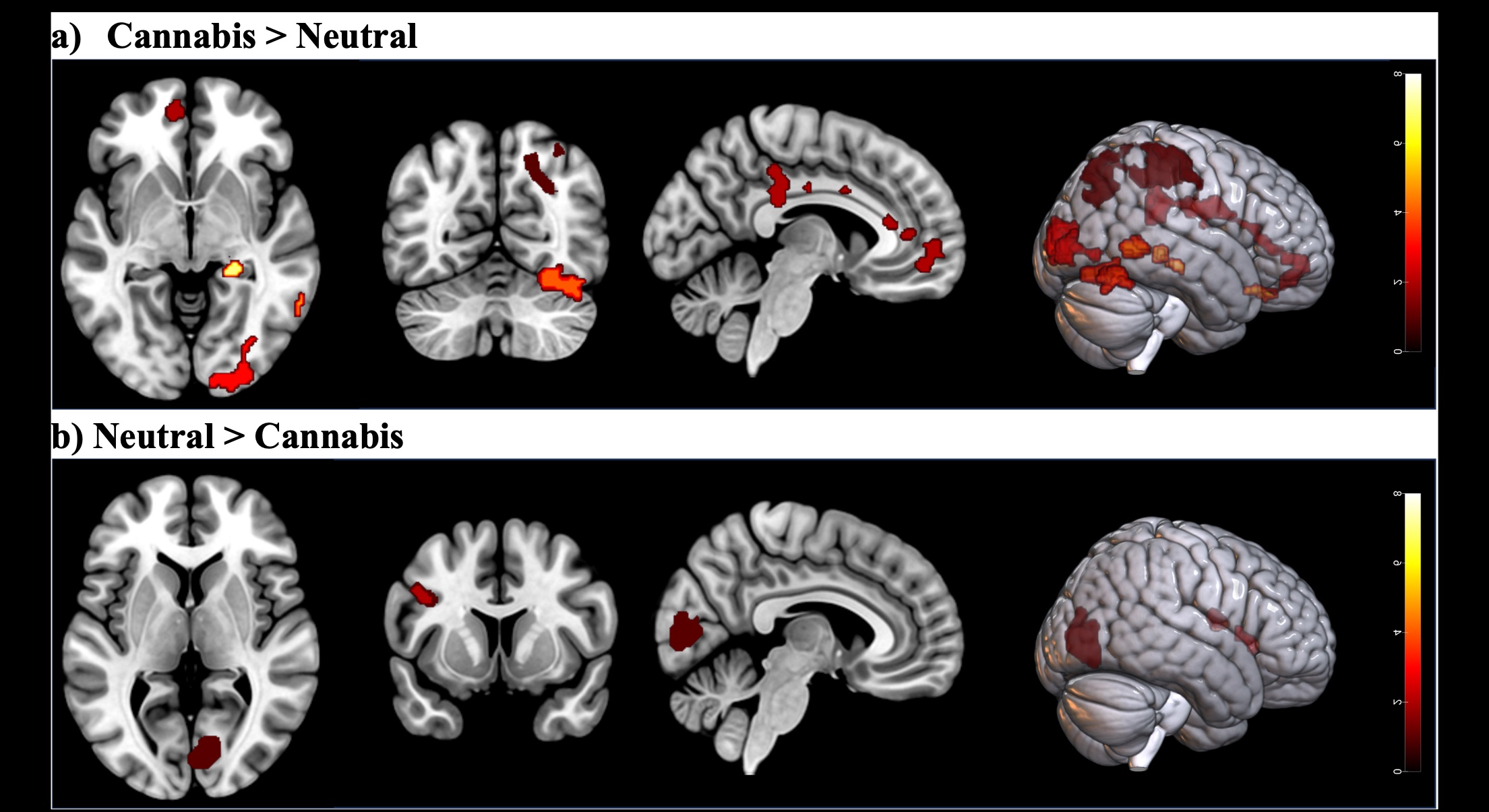
