## Supplemental Information for "The neurocircuitry of cue-induced cannabis craving in Cannabis Use Disorder: A functional neuroimaging study"

**Supplemental data**

1 Inclusion and exclusion criteria

**1.1 Inclusion criteria**

All participants:

i) age between 18 and 56 years;
ii) proficiency in English; and
iii) normal or corrected-to-normal vision.

Participants with a CUD:

i) endorsed DSM-5 criteria for a moderate-to-severe CUD based on the Structured Clinical Interview for DSM-5 Research Version (SCID-5-RV),
ii) consumed cannabis daily/almost daily for at least the past 12-months, and
iii) reported at least one attempt to quit or to reduce their use in the past 24 months.

**1.2 Exclusion criteria**

All participants:

i) significant medical conditions, history of acquired brain injury or loss of consciousness > 5 minutes;
ii) history of psychopathology (except for depression and anxiety) ascertained by the Mini International Neuropsychiatric Interview (MINI);
iii) illicit drug and alcohol use in the 12-hours before testing determined by self-report;
iv) significant alcohol use or dependence (i.e., Alcohol Use Disorder Identification Test (AUDIT (1); score ≥ 14);
v) illicit drug use (other than cannabis in the CUD group) in the past 4-weeks or above recreational levels (i.e., > 50 lifetime episodes, or > weekly use over a 3-month period) confirmed by a comprehensive questionnaire previously used by our team (2-4);
vi) current use of prescription medication that affects the central nervous system with the exception of antidepressants, e.g., selective serotonin reuptake inhibitors (SSRI’s) and serotonin-norepinephrine reuptake inhibitor (SNRI’s), due to increased prevalence of depression and anxiety in CUD populations and our inclusion of these mental health disorders (5);
vii) MRI contraindications (e.g., pacemaker, pregnancy) confirmed by a screener from the testing facility:	<https://www.monash.edu/__data/assets/pdf_file/0004/972022/pqms3-mbi-frm-c001-mri-screening-information-form.pdf>; and
viii) IQ scores < 80 determined by the vocabulary and matrix reasoning subtests of the Weschler Abbreviated Standardised Intelligence-II (6).
2 **Process of participants’ selection against inclusion and exclusion criteria**

A total of 9,030 people completed an the online screening survey against study’ inclusion and exclusion criteria. The measures used to address this included (1) demographics; (2) MRI screening; (3) previous research participation; (4) handedness; (5) past and current cannabis use related questions; (6) alcohol use history; (7) substance use history; (8) mental health screening; (9) depressive symptoms; (10) COVID-19 related stress and; (11) attempts to quit cannabis. Of these, 7,980 did not meet study eligibility or did not fully complete the screener. About ~1,050 people who were potentially eligible in the study underwent a phone follow up to confirm eligibility. Any queries about participants’ eligibility were resolved via discussion with the study CI and the research team, before confirming inclusion or exclusion.

A total of 117 participants aged 18-56 years (39 female) were included for face-to-face behavioural/MRI testing. This is where eligibility criteria were confirmed. The sample included one male participant in their 20s who has been using cannabis for 11 months (i.e., less than 12 month cannabis use history) who met eight DSM-5 criteria for a CUD (i.e., severe) and used daily. Nine participants were excluded after face-to-face testing when it emerged they met exclusion criteria.

**2.1 Reimbursement**

Participants were reimbursed via Coles/Myers vouchers of $100 for controls and $150 for the CUD group (additional reimbursement was to compensate for further research activities part of the broader study).

3 **Assessment of Sociodemographic, substance use and mental health data**

Questionnaires measured sociodemographic variables, including age and sex. Metrics of substance use and related problems and mental health.

#### 3.1 Metrics of cannabis exposure and related problems

The *Structured Clinical Interview for DSM-5 Research Version* (SCID-5-RV) was administered to determine the severity of CUD for inclusion and the number of symptom scores endorsed (7). The *Cannabis Withdrawal Scale* (CWS) was administered to measure withdrawal symptoms (8); while 10-point visual analogue scales (VAS) were given to rate participant’s ratings of *arousal and affective valence* of all the cannabis and neutral images that were viewed during the fMRI cue-induced craving task. Arousal was rated from 0 representing “*calm*” to 10 “*excited*”; and affective valence was rated on a scale from 0 “*unpleasant*”, to 10 “*pleasant*” with a rating of 5 representing “*neutral*”. A VAS scales was also administered to measure cannabis cravings immediately before and after the fMRI cue-induced craving task, with the item *“how much do you feel like smoking cannabis right now?”,* and answers ranging from ‘0’ indicating *“not at all”* and ‘10’ “*extremely”.*

A *semi-structured interview used* in previous studies was administered to measure lifetime cannabis use history, from which we extracted key cannabis exposure parameters accounting for periods of abstinence (2-4). They were: age when cannabis was first tried, age of regular cannabis use (i.e., defined as ≥ 2 days/month), duration of regular cannabis use (i.e., defined as at a rate of ≥ 2 days/month), and cumulative lifetime exposure in grams. The *Timeline Follow-Back* was administered to measure past month: days of use, dosage in grams, hours since cannabis was last use (9).

Presence of cannabis use was corroborated via presence of *11-nor-9-carboxy-Δ^9^-tetrahydrocannabinol:creatinine* (THC-COOH:creatinine) in urine, which measure THC concentration combined with creatinine which controls for the variation in THC due to varying hydration status of participants. It was confirmed by toxicology analyses conducted by Drugs and Toxicology Group, Centre for Forensic Science, University of Technology Sydney.

#### 3.2 Metrics of exposure to other substances and substance use-related problems

The *Timeline Follow-Back* was administered to extract days of use and dosage for alcohol (i.e., standard drinks) and nicotine use over the past 30 days (9). Alcohol and nicotine dependence were assessed via the *AUDIT* and the *Fagerström Test for Nicotine Dependence* (FTND; (10)).

#### 3.3 Mental health symptom scores

The *Beck’s Depression Inventory* – Second Edition (11)), the *State-Trait Anxiety Index – Y Form* (12), and the *Perceived Stress Scale* (13) were administered to measure depression, anxiety and perceived stress symptoms. The Community Assessment of Psychic Experiences (CAPE) was given to meaure positive and negative psychositc symptoms, and depressive symptoms (14). The *COVID-19 Stress Scales* were administered to measure COVID related stress in relation to: danger, socioeconomic consequences, xenophobia, contamination, traumatic stress, and compulsive checking (15).

### 4 MRI data acquisition

The neuroimaging data acquisition is outlined in detail in the following section and Supplemental Figure 1.

#### 4.1 Structural MRI

MRI images were acquired on a Siemens Skyra 3 Tesla scanner using a 32-channel head coil at the Monash Biomedical Imaging facility. Brain images were acquired in coronal view, from anterior to posterior. Structural MRI data was acquired using T1-weighted MPRAGE scan with the following acquisition parameters were: TE = 2.07ms, TR = 2300ms, flip angle = 9°, 192 slices without gap, FOV 256x256mm, 1x1x1 mm voxels, and total acquisition time of ~5 minutes and 20 seconds.

#### 4.2 fMRI cue-induced craving task

fMRI data during the cue-induced craving task was acquired using T2* weighted EPI scans. Acquisition parameters were: TR = 2240ms, TE = 30ms, flip angle = 90°, field of view = 192mm, matrix = 64, voxel size 3 x 3 x 3mm^3^, 40 slices, with 227 total volumes. The task total acquisition time was ~8 minutes and 37 seconds.

a. ***Task description***

An event-related cue-induced craving fMRI task adapted from previous experiments from our extended team (16) was used to measure brain activity while participants passively watched 30 cannabis and 30 neutral non-cannabis images used as cues of comparable resolution, type of activity, size, brightness, and luminance (Supplemental Figure 1).

The images showed cannabis-related paraphernalia and use behaviours, and neutral images depicted stationary items or cooking utensils. Each image was presented for 4 seconds preceded by a fixation-cross that lasted on average 4 seconds, jittered between 2 and 6 seconds. The cannabis and neutral images were presented in the same semi-random order (max three images of the same category in a row) for each participant.

Participants were instructed to pay close attention to the images “*In this task you will see pictures on the screen. Please try to keep your head still. Your task is to look at these pictures closely and as attentively as you can. The task will take about 10 minutes. We are about to start. Are you ready?*”. To ensure that participants maintained attention, their alertness was monitored via an MRI compatible camera and any occasions of sleepiness prompted a re-start of the scan. Images were presented onto a rear projection screen positioned behind the MRI scanner using E-prime3 software (Version 3.0 Build 3.0.3.80, E-Studio Build 3.0.3.82, Psychology Software Tools Inc.). Total task time was ~10 minutes.

**4.3 MRI data pre-processing**

fMRI data pre-processing steps were completed via fMRIPrep (version 20.2.3, ref): distortion correction, head motion correction, slice timing, co-registration with T1-weights images normalised to standard space (i.e., Montreal Neurological Institute [MNI] space), and smoothing with 6 mm Gaussian kernel. Image quality was assessed (i.e., motion, signal-noise ratio, artifacts) against conservative criteria validated for resting-state fMRI via review of Framewise Displacement (FD; indicator of motion, including data < 0.5 mm cutoff) (13). No participants were excluded after fMRI quality checks.

5 **Overview of participants excluded from analyses after testing**

Nine participants (6 with a CUD and 3 controls) were excluded due to subsequently meeting exclusion criteria when face-to-face testing. The people with a CUD who were excluded were: a female in their 20s with an IQ score < 80 (i.e., FSIQ-2 = 61), one male in their 20s endorsing a neurological disorder, three participants (two male in their 30s and one female in their 20s) with incidental findings determined by a neurologist that conducted comprehensive checks of MRI images, and a male aged 30s who used synthetic cannabis. The three controls excluded reported > 50 lifetime occasions of cannabis use. They were: i) a female aged in their 30s years, who reported ~4000 lifetime occasions over 13.5 years, ii) a female aged 50s years, who reported 176 lifetime occasions since their 20s, and one male in their 30s, who reported 420 lifetime occasions in 14-months and endorsed a history of a diagnosed psychiatric condition in adolescence.

6 **Occasional illicit substance use**

As outlined in Supplemental Table 1, 20 participants used illicit substances other than cannabis, of which 2 were controls. They used substances other than cannabis over the 4-week period prior to scanning, on a median of 1 occasion (range 1 – 6 occasions), for a median of 229 hours before testing (range 16 – 648 hours).

**Supplemental Table 1. Overview of participants’ occasional exposure to illicit substances within 4 weeks from testing**

**[Insert Supplemental Table 1]**

*Abbreviations*: MDMA, methylenedioxy-methylamphetamine; _, missing; mg, milligrams.

7 **Brain activity in the whole sample during cue-induced craving fMRI task**

We examined brain function in the whole sample during cue-induced craving fMRI task as ‘sanity check’ to ensure the task recruited the expected neurocircuitry for the contrasts cannabis>neutral and neutral>cannabis.

In the whole sample, there was greater brain activity while participants viewed *cannabis > neutral* images (Supplemental Figure 1 and Supplemental Table 2) in a range of frontal, temporo-parietal and visual cortical regions brain regions: superior frontal gyrus medial/dorsolateral, middle temporal/occipital gyrus, anterior cingulate & paracingulate gyri, angular gyrus, middle temporal gyrus, calcarine fissure and supramarginal gyrus. Meanwhile, for the contrast *neutral > cannabis*, the whole sample showed greater activity in additional areas: inferior/middle occipital gyrus, precentral gyrus, inferior frontal gyrus (i.e., pars triangularis and pars opercularis), supplementary motor area, and superior medial OFC.

**[Insert Supplemental Figure 1]**

**Supplemental Figure 1**. **Brain regions of differences for the whole sample during the contrasts (a) cannabis > neutral, and (b) neutral > cannabis images, *p* <0.001; FWE-corrected in axial, coronal and sagittal and 3D views)**

**Supplemental** **Table 2. Brain regions implicated in the fMRI cue-induced craving task across the whole sample (*p*<0.001, FWE corrected)**

**[Insert Supplemental Table 2]**

*Note:* Superior medial OFC superior frontal gyrus medial orbital, left.

*Abbreviations*: k, cluster size; *p*, p-value; FWE, family-wise error; FDR, family discovery rate; TFCE, threshold-free cluster enhancement; Z, z-statistic; unc, uncorrected; MNI, Montreal Neurological Institute; AAL, Automated Anatomical Atlas; ACC, anterior cingulate cortex; OFC, orbitofrontal cortex.

8 **Additional results on correlations before and after outlier removal**

**9.1 Descriptives for participants who met criteria for outliers**

Sensitivity brain-behavior correlations were run in the whole sample before and after outlier removal. Outliers are described below as a function of the relevant variable.

There were 6 outliers, for alcohol standard drinks/past month. They included 2 participants with moderate CUD (i.e., 206.8 drinks for a male aged in their 40s; 201 drinks for a female in their 20s); and 4 participants with severe CUD (i.e., 156.4 drinks for a female in their 20s; 150.9 drinks for a male in their 30s; 111.4 drinks for a male in their 20s; and 101.4 drinks for a male in their 30s).

Duration of abstinence from cannabis use showed 8 outliers. Of these, one endorsed a moderate CUD (i.e., 64.5 hours for a male in their 20s). The remainder had severe CUD and included 6 males aged in their 20s and 30s (i.e., 73.25 hours; 44 hours; 44 hours; 40.5 hours; 38.25 hours) and 2 females in their 20s (i.e., 38.25 hours; 48 hours). Outliers also included a male with moderate CUD in their 30s, with a BDI score of 46; and a female in their 20s with severe CUD, with β values for middle occipital gyrus being 2.60.

9.2 **Correlations that were significant before and after outlier removal**

Within the CUD group, the following correlations were significant after outlier removal. These included: positive correlations between greater superior occipital cortex activity and arousal ratings of cannabis (*minus*) neutral images (VAS scores; rho = 0.40, *p* < .05), and withdrawal (CWS scores; rho = 0.31, *p* < .05); negative correlations between THC-COOH:creatinine levels & activities of the ACC and inferior parietal cortices (rho = -0.33, *p* < .05 and rho = -0.33, *p* <.05, respectively).

9.3 **Correlations that were significant in the whole sample & that did not survive outlier removal**

In the whole group of participants with a CUD, the following correlations were significant in the whole sample, but did not survive outlier removal. There were significant negative correlations between lower THC-COOH:creatinine levels and greater postcentral/supramarginal activity (rho = -0.30, *p* < .05 and rho = -0.28, *p* <.05, respectively); and positive correlations between greater middle occipital/cerebellum activity and withdrawal (CWS; rho = 0.33, *p* < .05 and rho = 0.34, *p* <.05, respectively) and positive correlations between the calcarine sulcus, middle occipital gyrus, and superior occipital gyrus with depression symptoms (BDI; rho = 0.29, *p* < .05 and rho = 0.34, *p* <.05, rho = 0.30, *p* <.05 respectively). After outlier removal, none of the results were significant.
